# dPQL: a lossless distributed algorithm for generalized linear mixed model with application to privacy-preserving hospital profiling

**DOI:** 10.1101/2021.05.03.21256561

**Authors:** Chongliang Luo, Md. Nazmul Islam, Natalie E. Sheils, John Buresh, Yong Chen

## Abstract

Hospital profiling provides a quantitative comparison of health care providers for their quality of care regarding certain clinical outcomes. To implement hospital profiling, the generalized linear mixed model (GLMM) is usually used to fit clinical or administrative claims data, adjusting for the effects of covariates. For better generalizability, data across multiple hospitals, databases or networks are desired. However, due to the privacy regulation and the computation complexity of GLMM, a convenient distributed algorithm for hospital profiling is needed. In this paper, we develop a novel distributed Penalized Quasi Likelihood algorithm (dPQL) to fit GLMM, when only aggregated data, rather than the individual patient data are available across hospitals. The dPQL algorithm is based on a newly-developed distributed linear mixed model (DLMM) algorithm. This proposed dPQL algorithm is lossless, i.e. it obtains identical results as if the individual patient data are pooled from all hospitals. We demonstrate the usage of the dPQL algorithms by ranking 929 hospitals for COVID-19 mortality or referral to hospice in Asch, et al. 2020.

## 1. Introduction

Hospital profiling provides a quantitative comparison of health care providers for their quality of care regarding certain clinical outcomes. An objective comparison across a heterogeneous healthcare system is important for consumers, providers, and policymakers. Such profiling across multiple hospitals is usually conducted by analyzing clinical or administrative claims data with proper statistical models [1, 2]. For fair comparison, patient-level factors including demographics, pre-existing conditions, disease severity and hospital-level factors including hospital volume are controlled for. For example, in a recent article of hospital profiling for COVID-19 mortality [2], the authors ranked the performance of 929 hospitals after adjusting for the patients’ characteristics including age, sex, Elixhauser comorbidities, and insurance type, and hospital’s characteristics including number of beds, number of ICU beds, urban/ nonurban setting, geographic region, profit status, and academic affiliation.

Recent years have seen the development of statistical methodologies for the purpose of hospital profiling. A commonly used model is generalized linear mixed model (GLMM), which assumes common fixed-effects of covariates, e.g. patient- and hospital-level factors, and hospital-specific random effects, i.e. intercepts on the interested clinical outcome [1, 2, 3]. Based on the estimated fixed and random effects, the risk standardized event rates (RSER) can be calculated for each site. GLMM estimation, though complicated, could be obtained by methods such as Gaussian-Hermite approximation of the integrated likelihood, Monte-Carlo based approaches, and penalized quasi-likelihood (PQL) approach [4, 5].

As mentioned before, hospital profiling usually relies on analyzing clinical or administrative claims data. For example, in Drye, et al. 2012, the investigators studied the in-hospital and 30-Day mortality rate of acute myocardial infarction (AMI), heart failure (HF), and pneumonia for more than 3000 hospitals using CMS medicare claims data [6]. In Asch, et al. 2020, the authors study COVID-19 mortality or discharge to hospice in 929 hospitals using the UnitedHealth Group Clinical Discovery Portal [2]. In both of these investigations, the studies were based on an integrated database, where patient level data were pooled into a single dataset. A potential limitation of the previous investigations, as pointed out by the reviewers of our earlier paper [2], is on the generalizability of the findings, because our investigation was based on the database from a single insurance company.

Ideally, if datasets from different institutes could be shared, the profiling methods can be applied to a more general study population. However, it is often the case that these healthcare data are protected by privacy regulations and communicating individual patient data are difficult. To extend hospital profiling to cover a wider patient population, privacy-preserving distributed algorithms can be used. Specifically, when fitting GLMM, the distributed algorithm is expected to require only aggregated data from each hospital (possibly in a few iterations) but obtains close estimates of the parameters, and therefore RSERs. There are some existing efforts on developing distributed algorithms for fitting GLMM. For example, Zhu, et al. 2020 proposed a distributed algorithm based on Expectation–Maximization (EM) algorithm [8]. However, it is well known that the EM algorithm usually takes many iterations to converge. As a result, the distributed algorithm also requires many rounds of data communication between institutes.

In this paper, we aim to fill this important methodological gap by proposing a novel distributed algorithm to fit GLMM that is lossless (i.e. it obtains identical results as if the individual patient data are pooled from all hospitals), computationally stable, and only requires few rounds of communications of aggregated data across institutes. The algorithm is based on the PQL approach and a newly developed distributed algorithm for linear mixed model (LMM). This proposed distributed PQL algorithm (dPQL) is lossless, computationally stable, and communication-efficient (i.e., only requires few rounds of communications). We demonstrate the usage of the proposed dPQL algorithms by hospital profiling for COVID-19 mortality or referral to hospice [2].

## 2. Methods

### 2.1 Fitting GLMM via penalized quasi-likelihood

GLMM is an extension of GLM with random effects. We introduce notations of GLMM in the context of hospital profiling. Assume there are *K* hospitals with numbers of patients *n*_*i*_, and the total number of patients is *N* = ∑_*i*_ *n*_*i*_. For subject j at hospital *i*, we denote *y*_*ij*_ the outcome, *x*_*ij*_ the *p*-dimensional covariates with fixed effects *β*, and *u*_*i*_ the random effect (i.e. random intercept), *i* = 1, …, *K, j* = 1, …, *n*_*i*_. Conditional on the covariates 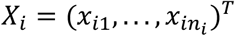 and random effects *u*_*i*_, 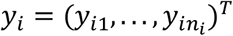 are assumed to be independent observations with means and variances specified by a GLM. Specifically,

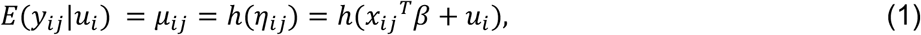

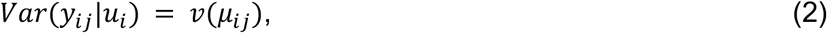

where *g* = *h*^−1^ is the link function that connects the conditional means *μ*_*ij*_ to the linear predictor *η*_*ij*_, *ν*(⋅) is the variance function. The random effects *u*_*i*_ are assumed to follow a normal distribution with mean 0 and variance θ.

Standard estimation of the GLMM parameters (*β, θ*) is based on maximizing the integrated quasi-likelihood

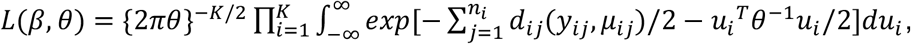

where

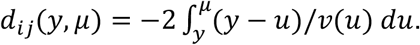

Maximization of this objective function is generally complicated [4], as the integrations must be performed numerically unless in the case of Gaussian outcome and identity link.

One approach to the integration is to make a Laplace approximation, which eventually leads to the PQL algorithm [4]. The PQL algorithm iteratively fit the linear mixed model

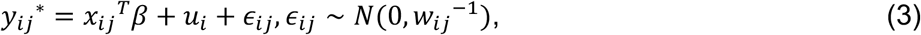

with the working outcome

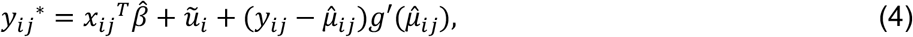

and the weight

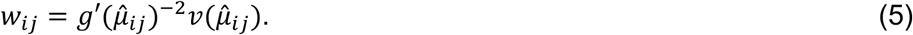

The obtained estimates are denoted as 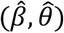. See [4, 5] for more details about the derivation.

### 2.2 The proposed dPQL algorithm

We develop a dPQL algorithm for GLMM estimation in the case that the individual patient data (IPD) are distributed across multiple centers and direct transfer of the IPD is not allowed. The dPQL algorithm is based on the distributed linear mixed model (DLMM) algorithm, which fits LMM exactly by requiring each site to contribute some aggregated data only once [9]. Specifically, in each iteration of the PQL algorithm, the weighted LMM (3) is fitted by the DLMM algorithm, requiring each site to contribute aggregated data

- *p* × *p* matrix *S*_*i*_ ^*X*^ = *X*_*i*_^*T*^ *W*_*i*_*X*_*i*_,
- *p − dim* vector *S*_*i*_ ^*Xy*^ = *X*_*i*_^*T*^ *W*_*i*_*y*_*i*_^*\**^, and
- scalars *s*_*i*_^*y*^ = *y*_*i*_^*\*T*^*W*_*i*_*y*_*i*_^*\**^, and sample size *n*_*i*_.

See the Appendix for details of the DLMM algorithm. The dPQL algorithm thus reconstructs the PQL iterations and obtains identical results as if the IPD are pooled together.

**The proposed dPQL algorithm**

1. Initialize: the lead site send an initial value of the fixed effects *β*^(0)^, and the random effects *u*_*i*_^(0)^ = 0 to the collaborative sites i=1, …, K.
2. For iteration s=0, 1, …,
  2.1 Site i calculates the working outcome *y*_*i*_^*\**^ = *η*_*i*_^(*s*)^ + (*y*_*ij*_ *− μ*_*i*_^(*s*)^)*g*′(*μ*_*i*_^(*s*)^), *η*_*i*_^(*s*)^ = *X*_*i*_*β*^(*s*)^ + *u*_*i*_^(*s*)^, and the weights *W*_*i*_ = *diag*{*g*′(*μ*_*i*_^(*s*)^)^−2^*ν*(*μ*_*i*_^(*s*)^)},
  2.2 Site i calculates aggregated data
    - *p* × *p* matrix: *S*_*i*_ ^*X*^ = *X*_*i*_^*T*^*W*_*i*_*X*_*i*_,
    - *p −* dim vector: *S*_*i*_ ^*Xy*^= *X*_*i*_^*T*^*W*_*i*_*y*_*i*_^*\**^ and
    - scalars: *s*_*i*_^*y*^ = *y*_*i*_^*\*T*^*W*_*i*_*y*_*i*_^*\**^and sample size *n*_*i*_, and transfers them to the lead site,
  2.3 The lead site fits weighted DLMM algorithm based on the aggregated data from 2.2, to obtain updated *β*^(*s*+1)^, *u*_*i*_^(*s*+1)^, and send them back to the collaborative sites.
3. Stop iteration when converged, e.g. || *η*^(*s*+1)^ *− η*^(*s*)^ || / || *η*^(*s*)^ || < 1e-6. The final estimates are 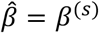, *ũ*_*i*_ = *u*_*i*_^(*s*)^ and 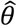

### 2.3 Distributed calculation for standardized mortality rates

We demonstrate that the standardized mortality rates of hospitals can also be calculated distributively without transferring the IPD, see Figure 1. We provide two approaches for risk standardization, the Standardized Mortality Rate (SMR) [1] and the Directly Standardized Mortality Rates (d-SMR) [3, 6]. While both approaches measure adjusted mortality rates effectively, d-SMR, in contrast to SMR, has an interpretation in an amenable probability scale.

**Figure 1.**
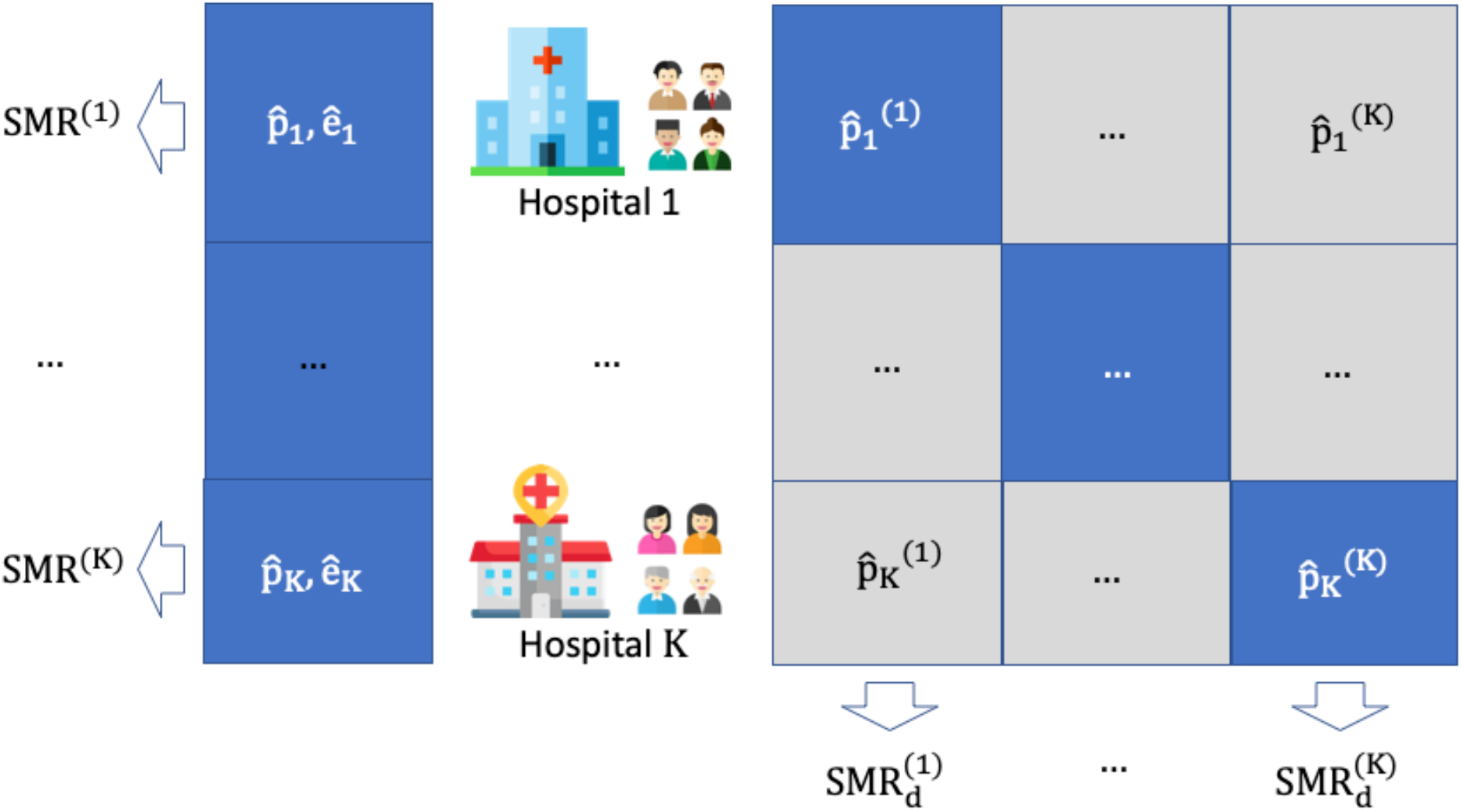
Standardized mortality rate of the hospitals can be calculated distributively based on the distributed PQL algorithm without transferring patient-level data across hospitals. On the left is the Standardized Mortality Rate (SMR), which requires hospital *k* to calculate its own average expected mortality rates 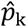 and *ê*_k_. On the right is the directly Standardized Mortality Rates (d-SMR), which requires hospital *i* to calculate and broadcast 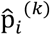, the average expected mortality rates of its patients had they been treated at hospital *k*. See equations (6-10) for computational details.

The SMR of hospital *k* is estimated [1] as

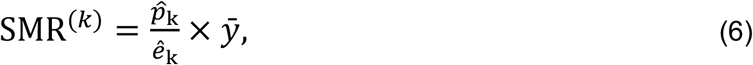

Where

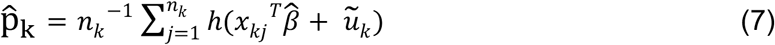

is the average expected mortality rate for patients at hospital *k*,

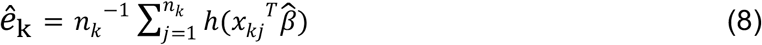

is the average expected mortality rate for hospital *k* patients had they been treated at the “population level,” and 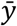 is the overall observed mortality rate. This SMR measure has been used to compare the performance of nonfederal acute care hospitals in the United States for acute myocardial infarction (AMI) (n=3,135 hospitals), heart failure (HF) (n=4,209 hospitals), and pneumonia (n=4,498 hospitals) from 2004 to 2006 [6].

The d-SMR of hospital *k* is defined as the average mortality rate assuming patients from all the hospitals being treated at this hospital [3], i.e.

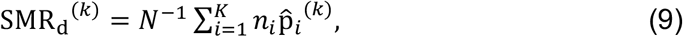

where

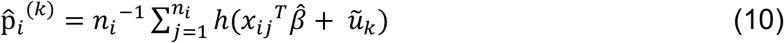

is the average predicted risk rates of patients at hospital *i* had they been treated at hospital *k*. When *i* = *k*, 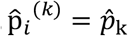, and if *i* = *k*, 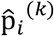 is a counterfactual probability. This d-SMR measure has been applied to profiling 4,289 hospitals in the United States for AMI using Medicare records from 2009 to 2011 [3], and to evaluating the change of COVID-19 mortality in 929 hospitals in the early period of the pandemic (January through April 2020) to the later period (May through June 2020) [2].

We note that with the proposed dPQL algorithm, both types of standardized mortality rate (SMR and d-SMR) measures can be calculated distributively without sharing patient level data. Specifically, for the SMR, hospital *k* calculates its own average expected mortality rates 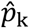 and *ê*_z_ using its own patient-level data and the public 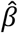 and *ũ*_k_ by equations (7-8), and thus SMR^(k)^ by (6). For the d-SMR, first, hospital *i* calculates and broadcasts 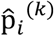 using its own patient-level data and the public 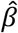 and *ũ*_k_ by (10), then each hospital *k* calculates 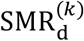 by (9). See Figure 1 for an illustration.

## 3. US hospital ranking based on the mortality rates for patients admitted with COVID-19

Asch et al., 2020 conducted a cohort study assessing 38,517 adults who were admitted with COVID-19 to 929 US hospitals from January 1, 2020, to June 30, 2020 using the data from UnitedHealth Group clinical discovery portal [2]. The hospital’s standardized rate of 30-day in-hospital mortality or referral to hospice was calculated, after adjusting for patient-level characteristics, including demographic data, comorbidities, community or nursing facility admission source, and time since January 1, 2020, hospital-level characteristics, including size, the number of intensive care unit beds, academic and profit status, hospital setting, and regional characteristics, including COVID-19 case burden; see Figure A1 for a description of the data.

We demonstrate the applicability of the proposed dPQL algorithm by applying it for ranking the hospitals with only transferring aggregated data from each hospital. Specifically, we compare the predicted mortality rate (via SMR or d-SMR) of the 929 hospitals by either pooled analysis (PQL) or the distributed analysis (dPQL) in Figure 2. The estimated fixed and random effects from dPQL algorithm and from the PQL are compared in Figure A2 in the Appendix. The dPQL algorithm reached convergence with only 5 iterations, and the estimation of fixed effects, best linear unbiased predictors (BLUPs), and mortality rates identical to that of the PQL from pooled data.

**Figure 2.**
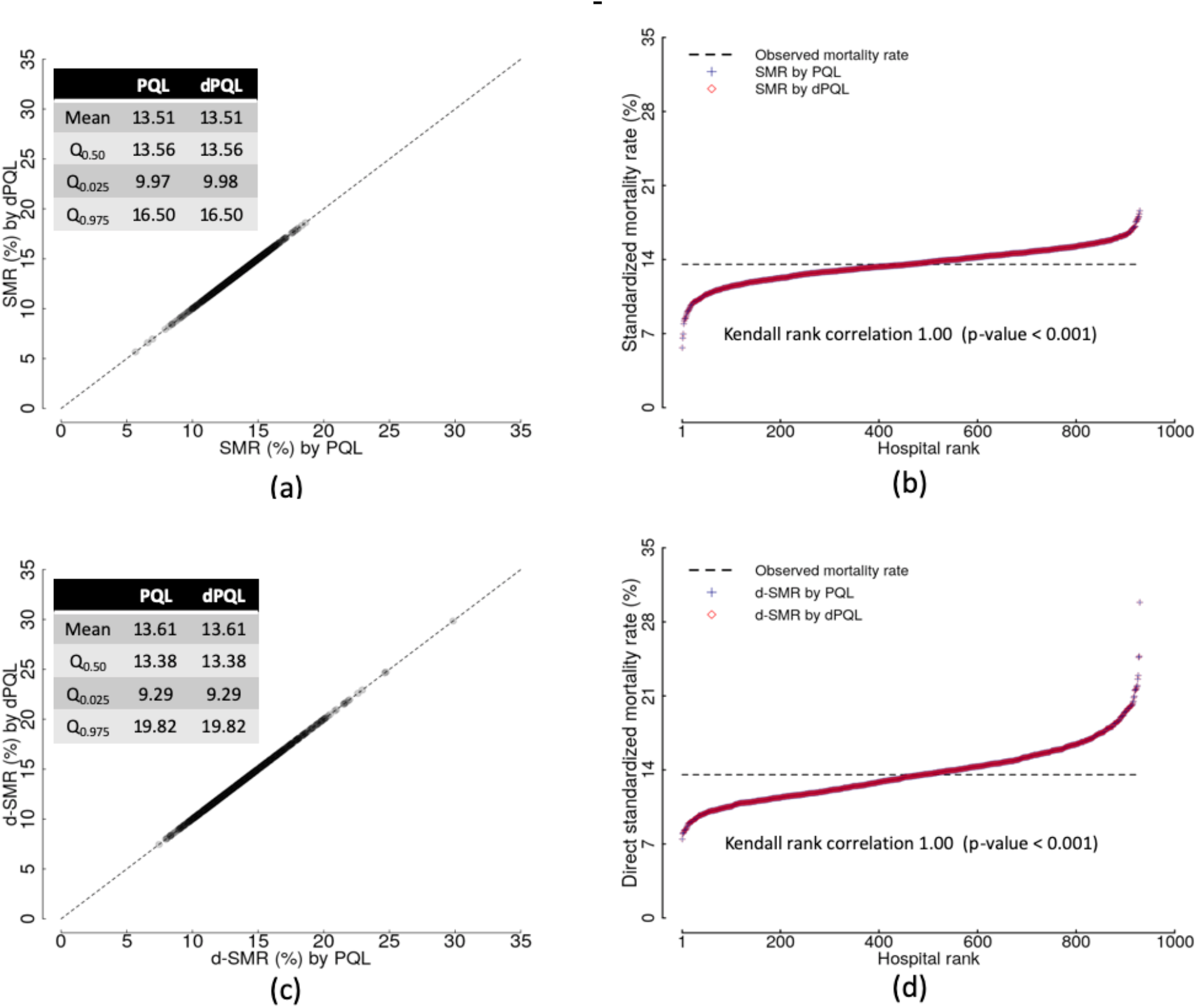
The estimated mortality rate (Standardized Mortality Rate, (a) and (b), and Directly Standardized Mortality Rates, (c) and (d)) of 30-day in-hospital mortality or referral to hospice of the 929 hospitals by either pooled analysis (PQL) or the distributed analysis (dPQL).

## 4. Discussion

We provide a privacy-preserving distributed learning algorithm, i.e. the dPQL algorithm to fit GLMM, which can not only obtain the identical results as if applying PQL method to the pooled data, but also conduct hospital profiling without sharing individual patient-level data. The proposed dPQL algorithm only requires sharing of minimal aggregated data from each site over few rounds of communications whilst obtaining identical results as if fitting GLMM to the pooled data using PQL.

The results of the PQL estimation are comparable to that of other approaches used to fit the GLMM model. For example, in the hospital ranking for COVID-19 mortality rates, the PQL estimation is almost identical to that of the Gaussian-Hermite approximation approach used in the original paper [2]. Although fitting GLMM by PQL is sometimes criticized for its biased estimation when the outcome is binary and clusters are small [4, 5], it is still an appropriate estimation approach for hospital profiling purposes, when the sample sizes in hospitals are large enough.

Since the PQL algorithm usually achieves convergence in a few iterations, we can consider a one-shot version of the dPQL algorithm, i.e. run only one iteration of the dPQL algorithm proposed in Section 2.2. This approach will sacrifice some accuracy of the estimation, but obtains very appealing communication cost, as each hospital need only to share the aggregated data once. Meanwhile, the number of iterations required in the PQL algorithm depends on the choice of initial values. While default initial values (i.e. all fixed effects being 0’s) provide satisfactory results, the performances can be improved with smart choices of initial values. Such strategy of further improving communication efficiency is currently under investigation, and will be reported in the future.

## Data Availability

The data are proprietary and are not available for public use but can be made available under a data use agreement to confirm the findings of the current study.

## 5. Acknowledgement

This work was supported partially through a Patient-Centered Outcomes Research Institute (PCORI) Project Program Award (ME-2019C3-18315). All statements in this report, including its findings and conclusions, are solely those of the authors and do not necessarily represent the views of the Patient-Centered Outcomes Research Institute (PCORI), its Board of Governors or Methodology Committee.

## Appendix

### The distributed linear mixed model (DLMM) algorithm

In each iteration of the PQL algorithm, it fits the linear mixed model *y*_*ij*_^*\**^ = *x*_*ij*_^*T*^*β* + *u*_*i*_ + *ϵ*_*ij*_, *ϵ*_*ij*_ ∼ *N*(0, *w*_*ij*_^−1^), with working outcome *y*_*ij*_^*^ and known weight 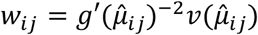. Below we show how this weighted linear mixed model can be fitted distributively by the DLMM algorithm.

The parameters in LMM are usually estimated by maximum likelihood (ML) or restricted maximum likelihood (REML) estimation [10]. Below we discuss the ML estimation and more details are in [9]. Since the random effects *u*_*i*_∼*N*(0, θ), the log-likelihood of LMM using all the data is

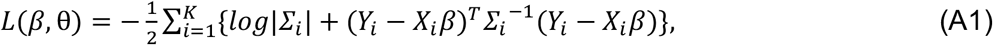

where *X*_*i*_ and 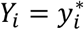 are the covariate matrix and the working outcome vector of the *i* ^th^ site respectively, |. | is the matrix determinant and 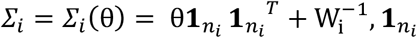 is a length *n*_*i*_ vector of all 1’s.

The maximum likelihood estimation can be further simplified by profiling out *β* from (A1). Given θ, the estimation of *β* is

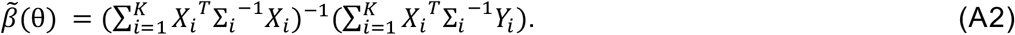

Thus the profile log-likelihood with respect to only θ is

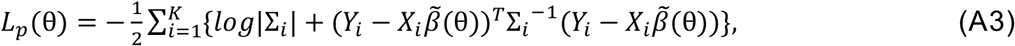

The ML estimate of θ can be obtained by maximizing (A3). The estimates of *β* can be subsequently obtained by (A2). We denote these estimates as 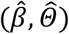. The variance of the estimated fixed effects 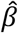 is thus

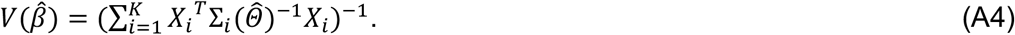

With some linear algebra, we can disentangle the data (*Y*_*i*_, *X*_*i*_) and the parameters θ in |Σ| and Σ_*i*_^−1^and thus reconstruct the profile log-likelihood (5) without communicating IPD. Specifically, we utilize the Woodbury matrix identity [11] to obtain

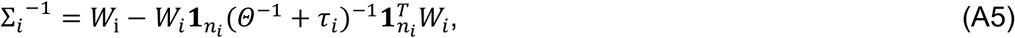

and the matrix determinant lemma [12] to obtain

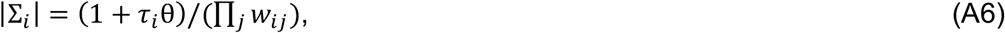

where *τ*_*i*_ = ∑_*j*_ *w*_*ij*_. The DLMM algorithm thus requires the *i*^*th*^ site to communicate

- *p* × *p matrix S*_*i*_ ^*X*^ = *X*_*i*_^*T*^*W*_*i*_*X*_*i*_,
- *p − dim vector S*_*i*_^*xy*^ = *X*_*i*_^*T*^*W*_*i*_*Y*_*i*_,
- *scalar s*_*i*_^*y*^ = *Y*_*i*_^*T*^*W*_*i*_*Y*_*i*_, *and sample size n*_*i*_,

for reconstructing the LMM likelihood. Specifically, to reconstruct (A3) with the above given aggregated data, we plug in (A5) to get (A2), then plug in (A2, A5, A6) to get (A3).

Finally, the BLUP [10] of the random effects *u*_*i*_ at the *i*^th^ site is

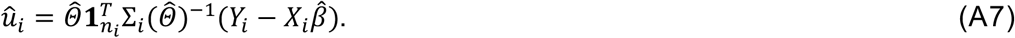

Conditioning on *X*_*i*_, *û*_*i*_ has mean zero and variance

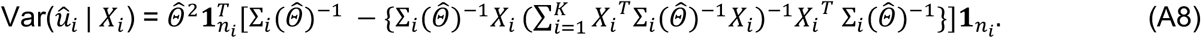

### US Hospital ranking for COVID-19 Mortality Rates

**Figure A1.**
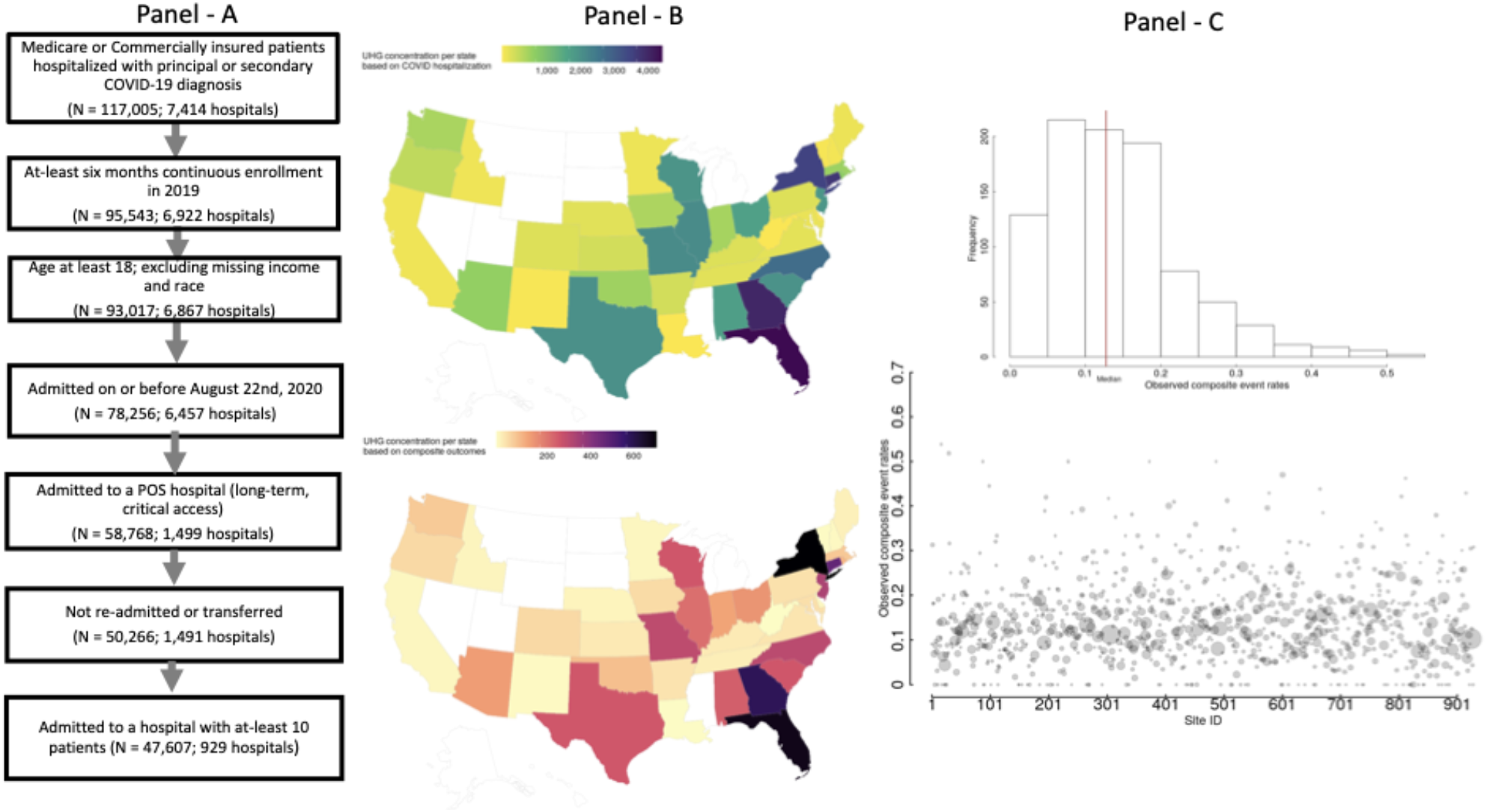
Data processing and summary of the hospital profiling.

**Figure A2.**
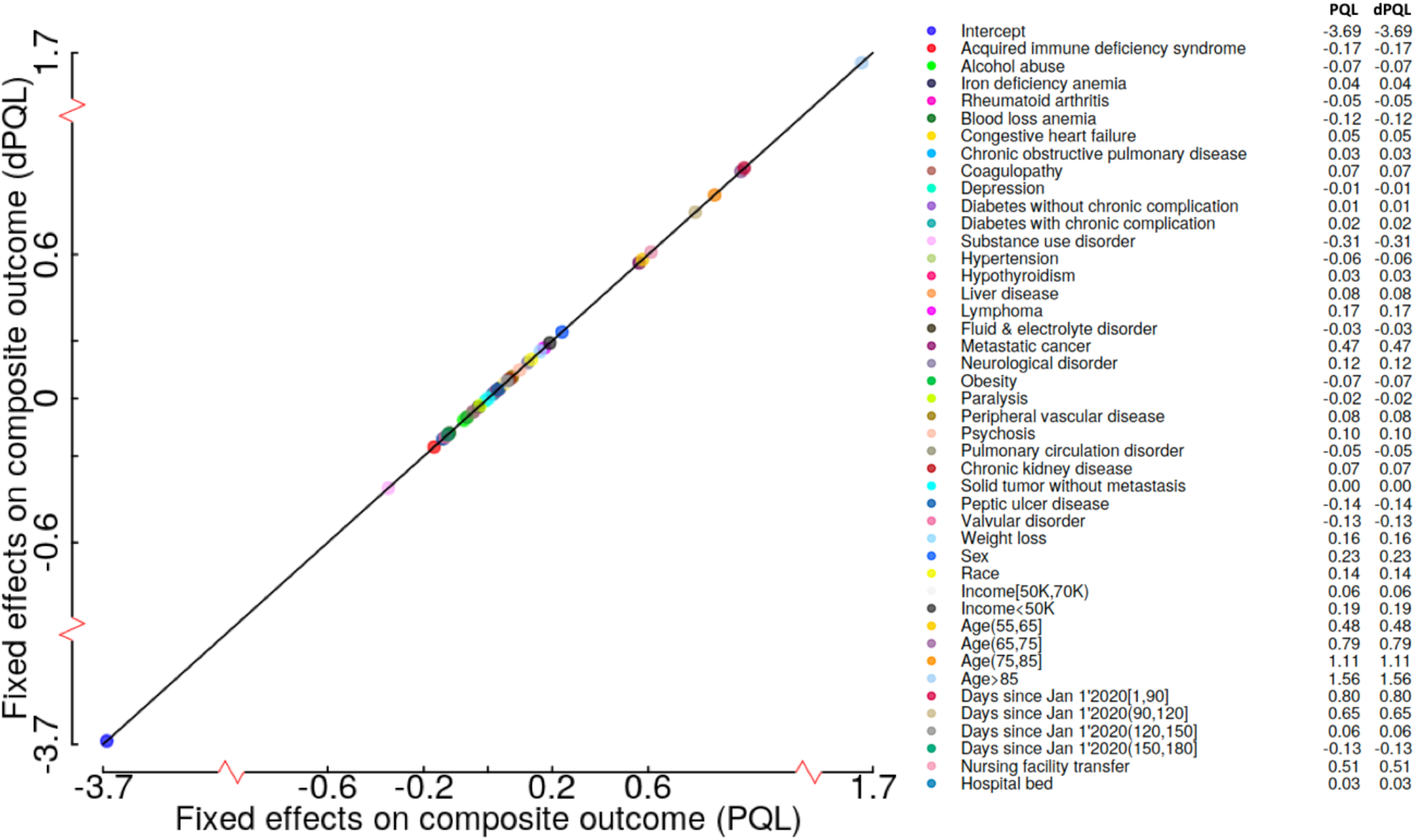
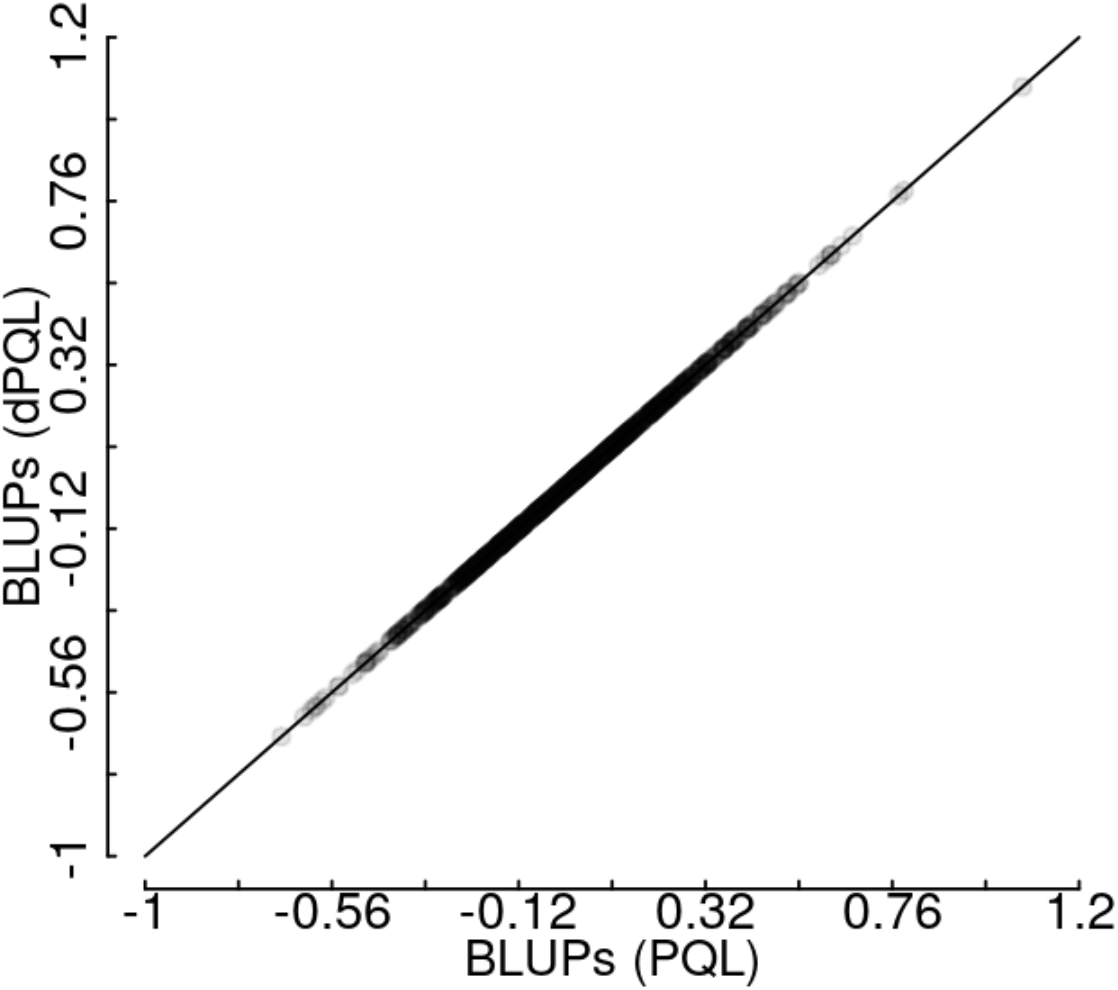
Comparison of the fixed effects and random effects estimation by the pooled analysis (PQL) and distributed analysis (dPQL) for ranking the hospitals regarding the COVID-19 mortality rate.

